# The St. Vincent’s Free Congestive Heart Failure Comprehensive Care Clinic: A Community-Based Intervention and Analysis

**DOI:** 10.1101/2022.12.23.22283823

**Authors:** John W Davis, Frederick S. Ditmars, Gabrielle Manno, Jacob Moran, Jenna Reisler, Elizabeth Davis, Khaled Chatila, Norman M Farr, Wissam Khalife, Robert D Thomas

## Abstract

**Introduction:** Heart Failure disease management clinics have been historically successful in reducing complications, but little has been done in uninsured settings.

**Methods:** This is a pilot program for uninsured HF patients following a recent hospitalization. Uninsured patients were offered enrollment in the disease management clinic during or immediately following hospitalization for a primary HF diagnosis at our institution during 2021. The program included twice-weekly visits with interprofessional support. Patients were scheduled 16 visits (2 months of follow-up) post-hospitalization. Patients who attended two visits were considered enrolled.

**Results:** Of 59 patients referred, 47(80%) were enrolled. Just four patients (8.5%,95%CI:2.5%,20.5%) were readmitted at 30 days, while four of twelve (33%,95%CI:13.6%,61.2%) were readmitted at 30 days in those who did not enroll. Program participants were readmitted significantly less frequently than national readmission rate estimates (23%,p=0.02).

**Conclusion:** The CHFC3 program is feasible and holds promise for materially reducing 30-day readmissions for HF complications in the uninsured.

## INTRODUCTION

Heart Failure (HF) hospitalization and readmissions remain a leading cause of morbidity, mortality, and cost-burden for the US healthcare system. As such, HF hospitalization has remained an intense focus of healthcare administration, especially since it holds strong ties related to reimbursement from the Center for Medicare and Medicaid Services (CMS). It is estimated that 35% of all 30-day readmissions reported to CMS are HF patients;^1^ of these, just 23% are for unavoidable reasons.^2^ Thus, current strategies have failed to substantially improve outcomes. HF prevalence is also rising, with an expected 772,000 more cases by 2040 in the United States alone.^3^ This is primarily related to an aging population trend; the average age at onset of HF occurs in the 8th decade of life (72-75 years of age for males and females respectively).^4^ Epidemiological surveillance data from previous decades^5^ suggest that prevalence of HF is less than 1% in persons younger than 50 years. Many of these patients are from low socioeconomic status (SES) backgrounds.^6,7^

National estimates for 30-day average readmission rates in HF range from 18-23%^1,8^ and are known to vary by age, sex, and overall illness burden. A nationally representative estimate using Healthcare Cost Utilization Project data demonstrates that 23% of all HF admissions are readmitted within 30 days, making HF the most common cause for 30-day readmissions of all diagnoses.^8^ This translates to patients being admitted with HF are automatically had a higher risk of readmission. Of approximately 6 million HF admissions from 2010-2017, 19.9% in 2017 alone were readmitted within 30 days.^1^

Interventions such as disease management clinics, nurse home visits, and nurse-care clinics are known to decrease HF readmissions.^9^ A network meta-analysis of 53 RCTs (n=12,356; mean ages 57-85) examined telephone visits, education sessions, pharmacist consultation, telemonitoring/support, nurse home visits, nurse case management, and disease management clinics compared to routine follow up for 30-day readmission.^9^ Nurse home visits significantly decreased HF readmissions by 35%,

Nurse Case Management decreased incidence by 23%, and Disease Management Clinics decreased rates by 20%. Other interventions were not significantly efficacious in reducing HF readmissions.^9^ Disease management clinics care typically included: follow-up with a cardiologist within two weeks of discharge, intermittent telephone consultation, and emphasis on clinical surveillance of vitals, medication adherence, and laboratory tests. However, patients in these studies were typically insured, older, and ethnically homogenous. Most of these interventions have been previously thought to be infeasible for those without coverage.

HF disparately affects patients who are uninsured or are socioeconomically disadvantaged. Of those in the United States who fall under the poverty level, approximately one-third also have HF.^10^ Being uninsured is commonly associated with other poverty markers including inability to afford transportation, housing, and food.

Poor healthcare access causes poor adherence to guideline-directed medical therapy, and thus more likely to suffer higher morbidity and mortality.^11^ In order to address this gap in care, social determinants of health (SDOH) such as job security, food security, and housing must also be considered. Therefore, the American Heart Association recommends an interprofessional approach aimed at addressing SDOH.^10^

Therefore, the purpose of this pilot is to assess the feasibility and potential effectiveness of a heart failure disease management program for the uninsured, which, to our knowledge, has not been previously attempted. Preliminary data from this intervention was for assessing a proposed multidisciplinary program at the St. Vincent’s Free Clinic (STVC) to prevent HF readmissions by providing free healthcare and free social support.

## METHODS

The design of this project is a single-intervention cohort without a control group (implementation analysis), though patients who did not enroll had limited information available for comparison. The study was determined to be exempt by UTMB’s IRB, as all patients received the same, free care if willing to enroll. This is a vulnerable population without reasonable alternative for care outside of the program, which precluded randomization since there are no alternative treatment options.

### Subjects

Inclusion criteria were identified by UTMB Cardiology staff from all UTMB hospitals prior to discharge. Patients who were: 1) uninsured, 2) admitted for a HF diagnosis-related group (either reduced or preserved ejection fraction) to a UTMB facility (as indicated by discharge note), and 3) willing to participate in the program were eligible for inclusion. Patients were informed that this follow-up program was designed to decrease complications in the immediate period following discharge and provides free medications and transportation to those who needed it.

Patients were discharged from January 4, 2021 to December 23, 2021, and subsequently offered enrollment in this program. Enrollment in the program was performed at the patient’s first appointment. Patients were identified by the St. Vincent’s staff as patients who were referred and in need of the program. All patients who attended at least the enrollment visit and a subsequent visit (at least two visits total) within 30 days post-discharge were considered enrolled.

### Procedure

The primary intervention was twice-weekly surveillance at the St. Vincent’s Clinic for vitals and medication adherence. Within 3 days of hospital discharge, patients were scheduled to receive care every Wednesday and Saturday at STVC over a period of 60 days, excluding holidays. Patients deemed “low risk” by their provider were eligible to decrease their visit frequency to once weekly after 30 days. Risk was determined according to each patients’ primary care provider’s judgment, but providers were encouraged to consider mortality risk models, such as the Seattle HF model,^12^ in making their decision. Patients were screened for SDOH needs and provided free medications, food, and transportation as indicated.

At Visit 1 (their baseline medical appointment following discharge), patients were queried on their interest in participation in CHFC3. Patients who declined were still offered care at STVC or connected to care elsewhere, according to preferences. All patients were given a full medical evaluation during Visit 1, where clinicians were instructed to initiate guideline-directed medical therapy (GDMT). All patients were given a standardized regimen of maximally-tolerated beta-blocker, SGLT2 inhibitor, mineralocorticoid receptor antagonist (spironolactone), and ACE/ARB/ARNI therapy as indicated. The program provided free medications to all patients by using social work support and charity funding. Patients’ vitals (blood pressure, heart rate, SpO2, weight, and respiration rate) were measured at each visit by a medical professional. Each week, patients also received a basic metabolic panel (BMP) measured to confirm renal function and electrolyte balances were unchanged, in addition to any other labs requested by faculty clinical staff. Abnormal lab values or vitals were reported directly to the supervising clinician. Patients were also connected to interprofessional services (Occupational Therapy, Respiratory Therapy, Nutrition, Pharmacist Consultation) as indicated. The templated schedule for interprofessional activities is demonstrated in **Table 1**.

**Table 1:** Weekly Activity Cadence.

| **WEEK/VISIT** | **ACTIVITY** |
| --- | --- |
| **1-1** | Medical evaluation with care provider, Vitals/Labs |
| **1-2** | Vitals review, medication adherence review |
| **2-1** | Occupational Therapy Initial Evaluation, Case Management (Social Work), Counseling (Psychologist), Vitals/Labs |
| **2-2** | Vitals review, medication adherence review |
| **3-1** | Pharmacist Consultation, Nutrition Consultation, Vitals/Labs |
| **3-2** | Vitals review, medication adherence review |
| **4-1** | Nurse visit, Vitals/Labs, Ad-Hoc Visits with other disciplines |
| **4-2** | Vitals review, medication adherence review |
| **5-1** | Medical evaluation (option for 'graduation if deemed medically appropriate'), Vitals/Labs |
| **5-2** | Vitals review, medication adherence review |
| **6-1 – 8-2** | Repeat from 2-1 to 4-2 |

During Week 2, patients received pharmacist consultation, counseling services, and occupational therapy evaluation. Case management (social services) was consulted to address outstanding social needs. Intermediate visits (Weeks 3-4) were performed by a nurse. Week 5 included an exit medical evaluation. In all visits, vitals and medication adherence were confirmed.

### Measures

The primary measures collected at Visit 1 in the program were age, sex, race/ethnicity, and discharging hospital. Secondary measures included basic metabolic panel (which included renal function and blood sodium), Brain Natriuretic Peptide (BNP), Vitals (Weight/BMI, blood pressure, heart rate, respiration rate), New York Heart Association (NYHA) functional class (I-IV), and history of diabetes mellitus (as measured by HbA1C), medications, and discharge ejection fraction. Patients who had a discharge ejection fraction > 45% (within normal limits) were defined as HF preserved Ejection Fraction (HFpEF), whereas those with <45% were defined as HF reduced Ejection Fraction (HFrEF). At their exit visit (after four weeks of program enrollment), patients who were still enrolled repeated BMP, BNP, Vitals, and NYHA Functional Class Assessment.

Patients were also queried about transportation and food insecurity, specifically as to whether transportation unavailability had historically precluded them from attending medical visits and whether they had ever had to go without food because of financial reasons. Lab draws were performed at regular intervals according to standard of care, and were unavailable where the clinician felt benefits were outweighed by risks of venipuncture. A table of all measures can be found in **Table 2**.

**Table 2:**
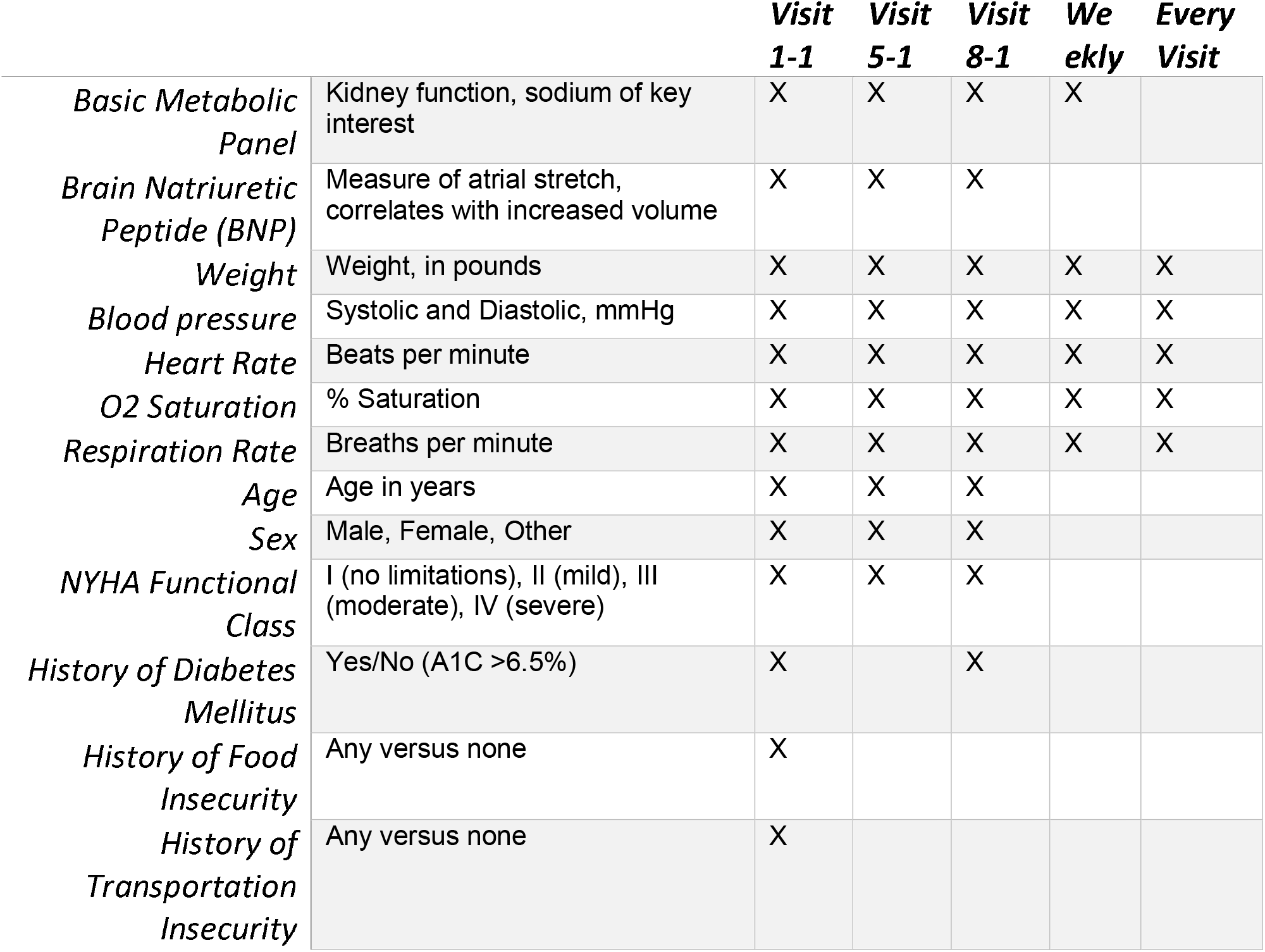
Key Measures.

| *Measure* | *Description* | *Visit*<br>*1-1* | *Visit*<br>*5-1* | *Visit*<br>*8-1* | *We*<br>*ekly* | *Every*<br>*Visit* |
| --- | --- | --- | --- | --- | --- | --- |
| *Basic Metabolic Panel* | Kidney function, sodium of key interest | X | X | X | X |  |
| *Brain Natriuretic Peptide (BNP)* | Measure of atrial stretch, correlates with increased volume | X | X | X |  |  |
| *Weight* | Weight, in pounds | X | X | X | X | X |
| *Blood pressure* | Systolic and Diastolic, mmHg | X | X | X | X | X |
| *Heart Rate* | Beats per minute | X | X | X | X | X |
| *O2 Saturation* | % Saturation | X | X | X | X | X |
| *Respiration Rate* | Breaths per minute | X | X | X | X | X |
| *Age* | Age in years | X | X | X |  |  |
| *Sex* | Male, Female, Other | X | X | X |  |  |
| *NYHA Functional Class* | I (no limitations), II (mild), III (moderate), IV (severe) | X | X | X |  |  |
| *History of Diabetes Mellitus* | Yes/No (A1C >6.5%) | X |  | X |  |  |
| *History of Food Insecurity* | Any versus none | X |  |  |  |  |
| *History of Transportation Insecurity* | Any versus none | X |  |  |  |  |

### Outcomes

#### Participation

First, a census of all uninsured patients admitted for HF, discharged, and subsequently referred to St. Vincent’s Clinic for care was identified using the medical record’s reporting tool. Patients who made at least one contact were subsequently identified. Participation was categorized as: 1) patients who never attended STVC (0 Visits), 2) patients who presented at STVC (1 Visit) but declined further participation, and 3) patients who completed at least two visits (which was considered enrollment). The total number of visits over the program course (within 2 months of discharge, up to 16 visits) for each patient was recorded. Participation was defined as at least two visits in the program.

#### Readmission

Patient history at 30 days after discharge were coded as readmitted (or died, with or without hospitalization) or not readmitted for any reason. Readmission location (UTMB or elsewhere) was recorded. Patients’ medical records were queried for any hospitalization or use of the emergency department, and findings were verbally confirmed with the patient at each visit. If patients missed their appointment, they were contacted to ascertain their admission status. Readmission was defined as any inpatient stay within 30 calendar days following the original discharge date (Day 0) to any location. All patients included in these analyses (enrolled or not enrolled) were followed from Index Date to Day 60.

### Statistical Analysis

The goal was to obtain pilot data on the feasibility and possible effect size of the CHFC3 Program in reducing all-cause readmission rates at 30 days in uninsured patients. In order to assess feasibility of participation, counts for program uptake (yes/no) and participation frequency (count of visits within 60 days of discharge) were obtained. 95% confidence intervals were estimated. Bivariate analysis of sociodemographic and medical history variables with participation measures was performed to assess selection bias. Possible selection bias magnitude was estimated by assessing sample differences by enrollment, using measures of association (phi for categorical variables such as diabetes history, and eta for continuous variables such as baseline age). Phi and eta are unbiased measures of association that are not sensitive to sample size, allowing for detection of possible selection bias even in small samples. T-tests and Mann-Whitney U testing was then performed to assess number of visits (0, 1, or 2+) by readmission. Visit counts were truncated at 2+ because patients who were readmitted were not able to attend the maximum number of visits (16), which would create survivorship bias in the analysis. Equal variance assumption was tested (alpha=0.10) for the t-test.

For estimating 30-day readmission rates in this program, the crude proportion of patients readmitted within 30 days who did not enroll (0 or 1 visit) versus those who enrolled (2+ visits) was compared using Fisher Exact Test. Subsequently, the readmission rate among those enrolled was compared to national estimates (23% by Fingar^8^, 19.9% by Khan^1^) using a two-sided, one-sample proportion test (alpha=0.05).

The pilot data generated from this program was used to estimate possible readmission reduction estimates when compared to national estimates. Previous evidence from interventions suggest that HF disease management clinics may be 20% effective^9^ in reducing HF readmissions. Power calculations were constructed for assessing the proportion of patients readmitted of those enrolled versus national readmission rate estimates. Assuming a reduction of 20%^9^ with CHFC3, compared to 19.9%^1^ (Khan) versus 23%^8^ (Fingar; intervention readmit rate of 15.9% or 18.4%, respectively at 30 days), at least 599 patients would be needed to achieve 80% power to detect this effect. Given this was a pilot program assessing feasibility, however, the aim here was to estimate the proportion of readmits that occurred and to estimate possible reduction in 30-day readmissions for future trials. All analysis was performed in SAS (Version 9.4, Cary NC).^13^

## RESULTS

### Cohort Description

From January 2021 to December 2021, there were 88 uninsured patients admitted to UTMB facilities (n=29 League City, n=59 Galveston) for a primary diagnosis of HF. Patients admitted to other campuses were not eligible for referral at program initiation; thus, just 2 of 29 (7%) non-Galveston based patients were referred to the program. Ultimately 61 (69%) total patients were referred to STVC for care, of whom 59 (97%) received at least one documented contact from the program. There are no data or information available for the 2 (3%) patients who were referred but were lost to follow-up.

Of those 59 patients referred and contacted, 47 (79.7%) completed at least two visits in the program, 3 (5.1%) attended just one visit, and 9 (15.3%) never attended any appointment. Overall, patients referred were a median of 53 years of age with a median 30% ejection fraction. 56% identified as non-White (either Hispanic or Black) and 34% were female. 51% were current smokers and while 34% reported frequent or daily alcohol intake. **Table 3** demonstrates the descriptive characteristics of patients by their enrollment status.

**Table 3:** Key Measures by Enrollment.

|  | **Not**<br>**Enrolled**<br>**(N=12)** | **Enrolled**<br>**(N=47)** | **Overall**<br>**(N=59)** | **Phi/Eta**<br>**Coefficient**<br>**(N=59)** |
| --- | --- | --- | --- | --- |
| **Age** |  |  |  |  |
| Median [Min, Max] | 54.5 [36.0,<br>65.0] | 52.0 [23.0,<br>78.0] | 53.0<br>[23.0,<br>78.0] | 0.03 |
| **Race/Ethnicity** |  |  |  |  |
| Non-Hispanic Black | 1 (8.3%) | 14 (29.8%) | 15<br>(25.4%) |  |
| Non-Hispanic White | 7 (58.3%) | 18 (38.3%) | 25<br>(42.4%) |  |
| White Hispanic | 3 (25.0%) | 15 (31.9%) | 18<br>(30.5%) |  |
| Unknown | 1 (8.3%) | 0 (0%) | 1 (1.7%) | 0.16 |
| **Sex** |  |  |  |  |
| Female | 6 (50.0%) | 14 (29.8%) | 20<br>(33.9%) | 0.17 |
| **BMI (kg)** |  |  |  |  |
| Median [Min, Max] | 31.0 [20.0,<br>67.8] | 29.9 [21.6,<br>66.7] | 29.9<br>[20.0,<br>67.8] | -0.09 |
| **NYHA Class** |  |  |  |  |
| Class I | 1 (8.3%) | 4 (8.5%) | 5 (8.5%) |  |
| Class II | 0 (0%) | 8 (17.0%) | 8 (13.6%) |  |
| Class III | 3 (25.0%) | 26 (55.3%) | 29<br>(49.2%) |  |
| Class IV | 6 (50.0%) | 9 (19.1%) | 15<br>(25.4%) |  |
| Unknown | 2 (16.7%) | 0 (0%) | 2 (3.4%) | -0.24 |
| **History of Diabetes Mellitus**<br>**(A1C >= 6.5%)** |  |  |  |  |
| Yes | 6 (50.0%) | 24 (51.1%) | 30<br>(50.8%) |  |
| Not screened | 2 (16.7%) | 0 (0%) | 2 (3.4%) | -0.07 |
| **Discharge Ejection Fraction** |  |  |  |  |
| Median [Min, Max] | 30.0 [15.0,<br>65.0] | 25.0 [10.0,<br>65.0] | 25.0<br>[10.0,<br>65.0] |  |
| Not acquired | 3 (25.0%) | 0 (0%) | 3 (5.1%) | 0.11 |
| **HFpEF** |  |  |  |  |
| Preserved EF | 3 (25.0%) | 7 (14.9%) | 10 (16.9%) | -0.11 |
| **Serum NT pro-BNP (mg/dL)** |  |  |  |  |
| Median [Min, Max] | 1440 [294, 6050] | 2410 [70.0, 18600] | 2220 [70.0, 18600] |  |
| Not acquired | 4 (33.3%) | 2 (4.3%) | 6 (10.2%) | 0.13 |
| **Creatinine (mg/dL)** |  |  |  |  |
| Median [Min, Max] | 1.08 [0.700, 1.98] | 1.18 [0.420, 3.86] | 1.17 [0.420, 3.86] |  |
| Not acquired | 3 (25.0%) | 1 (2.1%) | 4 (6.8%) | 0.09 |
| **History of Food Insecurity** |  |  |  |  |
| Yes | 4 (33.3%) | 21 (44.7%) | 25 (42.4%) | 0.09 |
| **History of Transportation Insecurity** |  |  |  |  |
| Yes | 4 (33.3%) | 15 (31.9%) | 19 (32.2%) | -0.01 |
| **Current Smoking** |  |  |  |  |
| Smoker | 7 (58.3%) | 23 (48.9%) | 30 (50.8%) | -0.08 |
| **Alcohol Consumption Frequency** |  |  |  |  |
| Multiple times per week or daily | 3 (25.0%) | 16 (34.0%) | 19 (32.2%) | 0.08 |

### Program Enrollment Measures and Baseline Characteristics

The mean number of visits attended by patients who enrolled was 8.3 (95% CI: 7.2, 9.4), and ranged from 2 to 16 visits. Program participants (n=47), despite being of younger age (median age 53 years) than the general HF population, had severe disease on average: the median discharge ejection fraction was 25%, 7 (14%) had HFpEF, and 74% of participants had Functional Class III-IV (at least symptoms at rest) on the New York Heart Association Disease scale. Participants also reported a high prevalence of important social determinants of health: 56% reported identifying as non-White (Hispanic or Black), 42% reported having food insecurity (missing meals regularly during the week due to finances), transportation insecurity (not having access to a vehicle or having missed an appointment because they did not have transportation at least once), 51% reported currently smoking, and 32% reported drinking alcohol multiple times per week to daily.

In the cohort who enrolled, the median Body Mass Index (BMI) was 30, 74% had NYHA Class III-IV, 7 (14%) had HFpEF, 51% had a history of diabetes mellitus, the median ejection fraction was 25%, and had a median creatinine of 1.18 mg/dL (mild to moderate renal disease). At baseline, HFrEF patients were prescribed guideline-directed medical therapy (GDMT): 63% were prescribed Sodium-Glucose Transporter 2 (SGLT2) inhibitors, 78% were prescribed an ACE, ARB, or ARNI drugs, 68% were prescribed mineralocorticoid receptor antagonists (spironolactone), 93% were prescribed a beta-blocker, and 70% were prescribed a statin. Only contraindications or patient refusal prevented clinicians from providing GDMT in this cohort.

### Readmissions Outcomes

The readmission rate, irrespective of program participation, was 13.6% (8 of 59, 95% CI: 6.8%, 24.8%). No patients died within 90 days post-discharge. Of the patients who enrolled, 8.5% (4/47, 95% CI: 2.5%, 20.5%) were readmitted within 30 days of discharge and 33.3% (4/12, 95% CI: 13.6%, 61.2%) were readmitted among those who did not enroll. 3 of the 4 readmissions in the non-enrolled group occurred in patients who attended 0 visits, whereas 2 of the 4 readmits in the enrolled group were in patients with preserved ejection fraction (2/7, 28.6%). Fisher’s Exact Test indicated that readmission rate significantly differed by enrollment (Phi=0.29, p=0.046). Readmissions also were significantly greater in unenrolled patients. Unadjusted odds of readmission were reduced by 81% (OR=0.19, 95% CI: 0.04, 0.90) in the enrolled versus unenrolled group.

It was hypothesized that disease severity may differ between enrolled and unenrolled patients, which would confound estimated treatment effect. he third analysis compared enrollment with readmission while controlling for disease severity. A simple severity index was created (NYHA Class IV or EF ≤15%). In the adjusted analysis, enrollment had a 79% reduction in odds of readmission at 30 days (OR=0.21, 95% CI: 0.04, 1.06). Although the effect was non-significant and the severity index had a large effect (OR=3.01, 95% CI: 0.33, 27.86). There was little change in the association between enrollment and readmission with (OR=0.21) and without (OR=0.19) adjustment for severity, suggesting that effect estimates were not influenced by HF severity differences.

The readmission rate of participants was significantly different from Fingar’s national average estimate (p=0.02), and Khan’s estimate (8.5% vs. 19.9%, p=0.050). However, since we did not have access to the raw data, it was not possible to assess whether sample differences confounded estimated rate differences.

## DISCUSSION

To our knowledge, this is the first programmatic attempt to connect uninsured HF patients to standard-of-care HF treatment – for free. This program was an implementation pilot project designed to replicate elements of prior, successful disease management programs. We found our program to have high feasibility and to be potentially effective in reducing HF readmissions, even when adjusting for disease severity and enrollment differences.

Compared to insured patients, uninsured patients are less likely to get care, are susceptible to poor care when it is received, and have overall worse health outcomes.^14^ What little care they are given is often disjointed, costly, inconvenient, and unsuccessful.^15^ Their poor health often limits employment potential, making it harder for them to obtain insurance, which further reduces likelihood of good health outcomes.^14^ This vicious cycle leads to a patient population that is at high risk for readmission as they are frequently lost to follow-up due to lack of access and/or inability to afford access.^16,17^ They do not have access to “routine” follow-up exams or the guideline-directed medical therapy shown to improve survival. Unfortunately, underinsured patients often have poor compliance due to their socioeconomic status.^18^ Food and housing instability were highly prevalent in our patient cohort, both of which have been associated with poor medication adherence.^18^ In our cohort, CHFC3 patients were substantially less likely to readmit at 30 days than national averages. They were also 80% less likely to be readmitted at 30 days than similarly discharged patients who did not enroll, which suggests possible effectiveness compared to similar, uninsured peers.

The program described here is comprehensive, and in some aspects, complex. It is also achievable and valuable. It helps bridge the gap that many underinsured patients experience by lacking a primary care provider.^19^ The program provides a reliable method of seeing a provider, obtaining medications, completing tests, and following with support services such as nursing, pharmacy, and therapy. During the study period, patients completed at least 8 visits within 60 days of discharge greater than 50% of the time; the majority of which occurred in the first month prior to discharge. It is thought that this intensive outpatient approach contributes to readmission prevention as readmission rates were significantly lower in the enrolled group versus the unenrolled (p=0.046) and the overall 30-day readmission rate was 8.5%. Enrolled patients’ readmission rate was significantly different from a nationally-representative readmission estimate (23% vs 8.5%).^8^

Hospitals may be unwilling to provide resources necessary for the conduction of this intensive surveillance study, which would limit generalizability. While the program included consultation with nurses, physicians, and other health professionals (i.e., nutritionists, pharmacists, etc.), these services are frequently unavailable for those who do not have means to pay. Further, consultation with these providers would require multiple visits in most other settings; STVC is unique nationally in providing all of these services all in one visit.^20^ However, it is important to be innovative in caring for the underserved, not only for patient quality and safety but also to reduce hospital costs. Given that each readmission prevented saves tens of thousands in direct costs to the institution,^21^ this program (if demonstrated effective in future studies) would be a cost-effective method of care.

Because patients with HFpEF were not excluded or studied separately in national readmission estimates, more work is needed to better appreciate whether this program holds promise in patients with preserved ejection fraction. Overall, 2 of 7 enrolled patients with HFpEF were readmitted within 30 days of discharge in this study. Because this sample is small, interpretation is limited. Just 5.0% (95% CI: 1.1%, 14.6%) of those with HFrEF were readmitted at 30 days. Further study is needed to evaluate whether the program is promising for those with HFpEF in addition to those with HFrEF.

### Limitations

Patients enrolled in this study were much younger than previously described cohorts (median age 53 years in CHFC3 versus 70+ in nationally representative cohorts),^22,23^ but generally still had advanced disease (75% of patients with NYHA Class >3, 50% with diabetes mellitus, 25% median ejection fraction). This is likely related to their low socioeconomic status which is associated with greater self-pay status and a greater prevalence of HF comorbidities.^17^ Nonetheless, findings from this pilot data should be interpreted with caution. This is further complicated by lack of a control group, precluding comparative effectiveness study. However, evaluation of patients who did not enroll allowed for crude estimations of possible selection bias. While this limited comparison did not reveal meaningful differences in baseline demographics and characteristics, more robust study methods are needed to assess possible program effectiveness.

Because this study does not have a control group, effectiveness cannot be estimated. However, the observed rates of participation and 30-day readmissions informs feasibility and rationale for future studies. There also may be selection bias in who chooses to attend CHFC3 versus those who do not enroll. While the estimated readmission rate was 8.5%, the global readmission rate for patients in this program was 13.6%. If all eligible patients had enrolled, this would suggest only a 32% reduction in readmission rate versus gold standard estimates, instead of the 57% reduction observed in the program. It is unclear what proportion, if any, of these readmissions would have been avoided if the patients had enrolled in the program.

COVID-19, which began in March 2020 in Texas and became exponentially more prevalent in the time since, likely affected the overall number of admissions and readmissions observed during this study. However, literature on the incidence of these outcomes during COVID-19 is limited. One retrospective cohort study in Philadelphia, USA, comparing HF admissions in a single urban hospital from March 2019-October 2019 versus March 2020-October 2020 indicated that HF hospitalizations overall decreased by 12% (p<0.001), but readmissions increased over time (19.1% vs 20.6%, p<0.001).^24^ However, internal UTMB data (unpublished) indicate 2020-2021 readmission rates in HF remained approximately constant at 19.8% (262/1326, 95% CI: 17.6%, 21.9%) versus the 19.9% observed from 2017-2019. Therefore, there is not sufficient evidence to suggest whether readmission rates differed because of COVID-19 in this study. Further, none of these patients were admitted for or received care for COVID-19 during the course of their program enrollment or previous hospitalization. Thus, it is unclear what if any effect COVID-19 had on this program.

While the program appears promising, some patients who did not enroll may have been more likely to readmit than those who chose to participate. Patients who enroll may be more motivated to remain adherent to medications, be less sick, or have less meaningful socioeconomic limitations than those who do not. However, association estimates between key, baseline measures (i.e., age, ejection fraction, NYHA Functional Class) and enrollment status were largely unremarkable. Thus, the potential for selection bias from anticipated confounders appears low, but a greater sample size in a more robust study design is warranted to appreciate the program’s true effectiveness.

## CONCLUSION

The CHFC3 program is feasible and holds promise for reducing readmissions in uninsured HF patients. It suggests that an intensive outpatient, multidisciplinary program may improve readmission rates in the underinsured, a marginalized and vulnerable group. The findings here warrant further exploration in clinical trials.

## Data Availability

Data is encrypted and stored in REDCap. This can be made available in de-identified fashion with confirmation from the University of Texas Medical Branch IRB.

